# Examining Differences in the Genetic and Functional Architecture of ADHD Diagnosed in Childhood and Adulthood

**DOI:** 10.1101/2023.08.02.23293439

**Authors:** Sophie Breunig, Jeremy M. Lawrence, Isabelle F. Foote, Hannah J. Gebhardt, Erik G. Willcutt, Andrew D. Grotzinger

## Abstract

**Background:** Attention-deficit hyperactivity disorder (ADHD) is a neurodevelopmental disorder that is diagnosed when symptoms onset in childhood. Here we investigate whether individuals who are diagnosed as children differ from those who are diagnosed in adulthood with respect to shared and unique architecture at the genome-wide, functional, and gene expression level of analysis.

**Methods:** We utilize Genomic SEM to investigate the differences in genetic correlations of childhood and adulthood diagnosed ADHD with 98 behavioral, psychiatric, cognitive, and health outcomes. We go on to apply Stratified Genomic SEM and transcriptome-wide SEM (T-SEM) to identify functional annotations and patterns of gene expression associated with genetic risk sharing or divergence across the ADHD subgroups.

**Results:** Relative to the childhood subgroup, adulthood diagnosed ADHD exhibited a significantly larger negative *r_g_* with educational attainment, the noncognitive skills of educational attainment, and age at first sexual intercourse. We also observe a larger positive *r_g_* for adulthood diagnosed ADHD with major depression, suicidal ideation, loneliness, and a latent internalizing factor. At the functional and gene expression levels, Stratified Genomic SEM and T-SEM analyses revealed three annotations and 22 genes, respectively, that were significantly associated with genetic risk sharing across the subtypes.

**Conclusion:** This project demonstrates that ADHD diagnosed later in life shows much stronger genetic overlap with internalizing disorders and related traits, indicating the potential clinical relevance of distinguishing these subgroups. Given shared clinical features across internalizing disorders and ADHD (e.g., difficulty concentrating) one interpretation is that the internalizing spectrum is often misdiagnosed as ADHD in adulthood.

## INTRODUCTION

ADHD is a neurodevelopmental disorder that affects ∼5% of the population and is characterized by a persistent pattern of inattention and/or hyperactivity and impulsivity^1^. Although an ADHD diagnosis can be given at any age, symptoms must onset before the age of 12 to meet diagnostic criteria. Given symptoms with approximately equivalent onset in childhood, age at ADHD diagnosis should theoretically not reflect a relevant clinical distinction. Yet, whether ADHD diagnosed later in life is differentially associated with outcomes and predictors remains a largely open question. For a disorder estimated to be ∼80% heritable^2^, genetic tools offer an exciting opportunity to better understand whether age at diagnosis reflects a relevant clinical and etiological specifier.

Multiple lines of evidence support a model of genetic divergence for ADHD diagnosed at different points in development. For example, ADHD diagnosed in childhood may reflect symptoms that are more severe and thereby clinically detectable at a younger age. The prediction here might be greater genetic overlap with childhood ADHD and clinical markers of severity (e.g., suicide attempts). Poor retrospective recall of childhood symptoms in adulthood^3^ may also result in genetic divergence due to consequent misdiagnosis. Indeed, recall bias coupled with shared symptoms across ADHD and other psychiatric disorders has led others to hypothesize that misdiagnosis is likely to be higher for ADHD diagnosed in adulthood.^4^ If this misdiagnosis in adulthood were particularly skewed toward one class of disorders the result would be higher levels of estimated genetic overlap with these same disorders.

The current study applied multivariate genomic tools to examine whether age at ADHD diagnosis demarcates different etiological boundaries and clinical distinctions at multiple levels of analyses. At the genome-wide level, we compared genetic overlap across child and adult diagnosed ADHD with psychiatric, cognitive, health, social, and behavioral outcomes. At the functional and gene expression level, we examined whether different classes of genetic variants (e.g., evolutionarily conserved) or patterns of gene expressions are associated with genetic risk sharing or uniqueness across subgroups. These findings collectively offer critical insight into the shared and unique genetic architecture across different ages of diagnosis.

## METHODS AND MATERIALS

### Phenotype selection

#### ADHD Stratified by Age at Diagnosis

The childhood and adulthood diagnosis ADHD GWAS were taken from the original paper by Rajagopal et al. and consist of a Danish ADHD case/control sample generated by iPSYCH. This sample included 14,878 childhood ADHD cases, 6,961 adulthood ADHD cases, and 38,303 controls that were randomly selected from the same nationwide birth cohort.^5^ The persistent ADHD phenotype (1,473 cases), reflecting individuals diagnosed before 18 that continued to show symptoms in adulthood, was excluded from the primary analyses as the *Z*-statistic for the SNP-based heritabilities was below the recommended cut-off for producing interpretable genetic correlation estimates^6^. The same set of controls was used for all subgroup ADHD GWAS to maintain a consistent reference point across subgroups.

Individuals were defined as ADHD cases if they had an ADHD diagnosis according to the ICD-10 criteria (F90.0 diagnosis code). Individuals were defined as being diagnosed in childhood if they were under the age of 18 at the time of diagnosis and as adulthood diagnosed if they were 18 years or older. Persistent ADHD was excluded from the analyses because the genetic correlations (*r_g_*) with childhood and adulthood diagnosed ADHD were high enough that we would have lacked the power to detect a genetic difference. Information on the genotyping, QC and GWAS procedures of the ADHD dataset can be found in the original article by Rajagopal et al.^5^.

#### External Traits

We selected a broad range of phenotypes to examine patterns of genetic overlap with childhood and adulthood diagnosed ADHD from the six broad domains of: psychiatric, cognitive, health, risk taking, social relationship, and substance use outcomes. For each phenotype, we selected the most well-powered, publicly available GWAS of unrelated individuals of European ancestry. A comprehensive list of the traits, including information such as GWAS sample size, can be found in Table S1. Details about dataset quality control can also be found in the **Online Supplement.**

### Genomic Structural Equation Modeling

We applied Genomic SEM to identify genetic correlations with external traits that were significantly different between the ADHD subgroups. This was achieved by first estimating the genetic correlations across traits using a multivariable version of LD-score regression^6^. We then estimated models in which the genetic correlation across the ADHD subgroups was freely estimated while the genetic correlations with an external trait were fixed to equality across the subgroups (**Fig. S1**). This produced a model with 1 degree of freedom (*df*), such that the model χ^2^ for this model specification reflects the level of model misfit resulting from the equality constraint. Thus, significant model χ^2^ estimates from this model represent external traits with significantly different levels of genetic overlap across the ADHD subgroups. A strict Bonferroni correction was used to correct for multiple testing by dividing the standard significance threshold of *p <* .05 by 98 traits, yielding a significance threshold of *p* < 5.10 × 10^-4^.

The primary analyses presented in the main text reflect the model χ^2^ estimates when using the genetic correlation and sampling correlation matrix (the standardized matrix of sampling dependencies) as input. Results using unstandardized estimates with the genetic covariance and sampling covariance matrix as input, can be found in Table S3.

### Internalizing Factor Follow-up Model

Many external traits that exhibited significantly stronger genetic correlations with adulthood diagnosed ADHD reflect traits in the internalizing domain. Given this trend, we conducted a set of follow-up analyses that included an internalizing factor defined by major depressive disorder (MDD)^7^, anxiety (ANX)^8^, and post-traumatic stress disorder (PTSD)^9^. We used this internalizing factor to first confirm that the pattern of larger genetic overlap with adulthood diagnosed ADHD we observe for individual internalizing traits also holds for this latent internalizing factor (**Fig. S2**). To further clarify this one set of results we ran two follow-up models testing whether the genetic correlation between childhood or adulthood diagnosed ADHD with internalizing disorders could be constrained to be equal to that observed between persistent ADHD and internalizing.

We went on to examine whether the significant differences between childhood and adulthood diagnosed ADHD with other external traits is accounted for by the genetic overlap between adulthood diagnosed ADHD and the internalizing space. To this end, we specified a model where this same internalizing factor predicted both childhood and adulthood diagnosed ADHD and the external traits to estimate differences in genetic overlap (applying a Bonferroni correction) when removing shared variance with the internalizing factor (**Fig. S3**).

### Stratified Genomic SEM

We utilized Stratified Genomic SEM^10^ to identify differential enrichment within functional annotations for childhood and adulthood diagnosed ADHD as well as the functional annotations that are common to both. Functional annotations refer to genetic variants grouped together based on some shared characteristic, such as the tissue types or neuronal subtypes where the variants are expressed. We began by running multivariable Stratified LDSC (S-LDSC)^11^ to obtain genetic covariance estimates within each annotation. We specifically included 55 annotations from the 1000 Genomes Phase 3 BaselineLD Version 2.2^12^, along with neuronal and brain tissue annotations from GTEx^13^, DEPICT^14^, gnomAD^15^, and Roadmap^16^. We then used Genomic SEM’s *enrich* function to estimate enrichment at the level of a general ADHD factor defined by both subgroups and for the residual genetic variance unique to childhood and adulthood diagnosed ADHD. The loadings on the ADHD factor were constrained to be equal to ensure model identification. We applied a Bonferroni corrected significance threshold for 172 annotations * three phenotypes (childhood diagnosed ADHD, adulthood diagnosed ADHD and the factor of both) of *p* < 9.69 × 10^-5^.

### Transcriptome-wide SEM

Transcriptome-wide SEM (T-SEM)^17^ was used to identify genes whose expression was associated with genetic risk sharing or uniqueness across the ADHD subgroups. We began by applying FUSION^18^ to estimate univariate TWAS for each subgroup. FUSION imputes the relationship between gene expression and a trait of interest using TWAS weights that reflect the associations between genotypes and gene expression levels from an external sample. We specifically included 16 sets of weights reflecting (*i*) 13 brain tissue weights from the Genotype-Tissue Expression project (GTEx v8)^19^, (*ii*) two dorsolateral prefrontal cortex weights from the CommonMind Consortium (CMC) ^20^, and (*iii*) one set of prefrontal cortex weights from PsychEncode.^21^ This resulted in 73,839 expression-imputed genes across the different tissues.

The gene expression estimates from the TWAS output were then combined with the LDSC covariance matrix to estimate the effect of gene expression on a general ADHD factor. In addition, we estimated the Q_Gene_ heterogeneity statistic, which pulls out genes that do not conform to the factor model (**Fig. S4**). In the context of the current analyses, this identifies genes whose expression is likely to be unique to either child or adult diagnosed ADHD. Hits for Q_Gene_ were defined using the Bonferroni corrected threshold of *p* < 6.77 × 10^-7^. Hits on the ADHD factor were defined using the same significance threshold and additionally excluded any Q_Gene_ hits. We also report univariate TWAS results for adulthood and childhood diagnosed ADHD separately using the same Bonferroni corrected significance threshold. We additionally conducted follow-up analyses for gene sets significantly associated with the ADHD factor using an over-representation analysis performed with the WebGestalt R package. However, these analyses did not identify any significantly overlapping gene sets.

## RESULTS

### Genomic SEM Reveals Divergent Genetic Correlations

The genetic correlation (*r_g_*) across childhood and adulthood diagnosed ADHD was 0.76 (*SE =* 0.06), which was significantly different from 0 and 1, thereby indicating both shared and unique genetic architecture across these subgroups. Consistent with this shared genetic architecture, we observed several external traits that were sizably and significantly associated with both subgroups, including migraines, aggression, and smoking outcomes (**Fig. S5**; also see **Fig. S6** and **S7** for depiction of top 15 genetic correlations with adult and childhood diagnosed ADHD, respectively). In addition, we did not observe significant differences for medical, substance use traits or circadian rhythms (**Fig. S8**).

In line with genetic divergence across these subgroups, we identified seven external traits with significantly different genetic correlations for childhood and adulthood diagnosed ADHD (**Fig. 1**; standardized results in Table S2; unstandardized results in Table S3). These seven traits included the aggregate ADHD GWAS, which uses both childhood and adulthood diagnosed subgroups. We found that the genetic signal for this combined ADHD GWAS most strongly overlapped with childhood diagnosed ADHD (*r*_g_Child_ = 0.95 [*SE* = 0.05]) relative to adulthood diagnosed (*r_g_Adult_* = 0.87; [*SE* = 0.06]; *p*_difference_ = 4.22 × 10^-4^).

**Figure 1.**
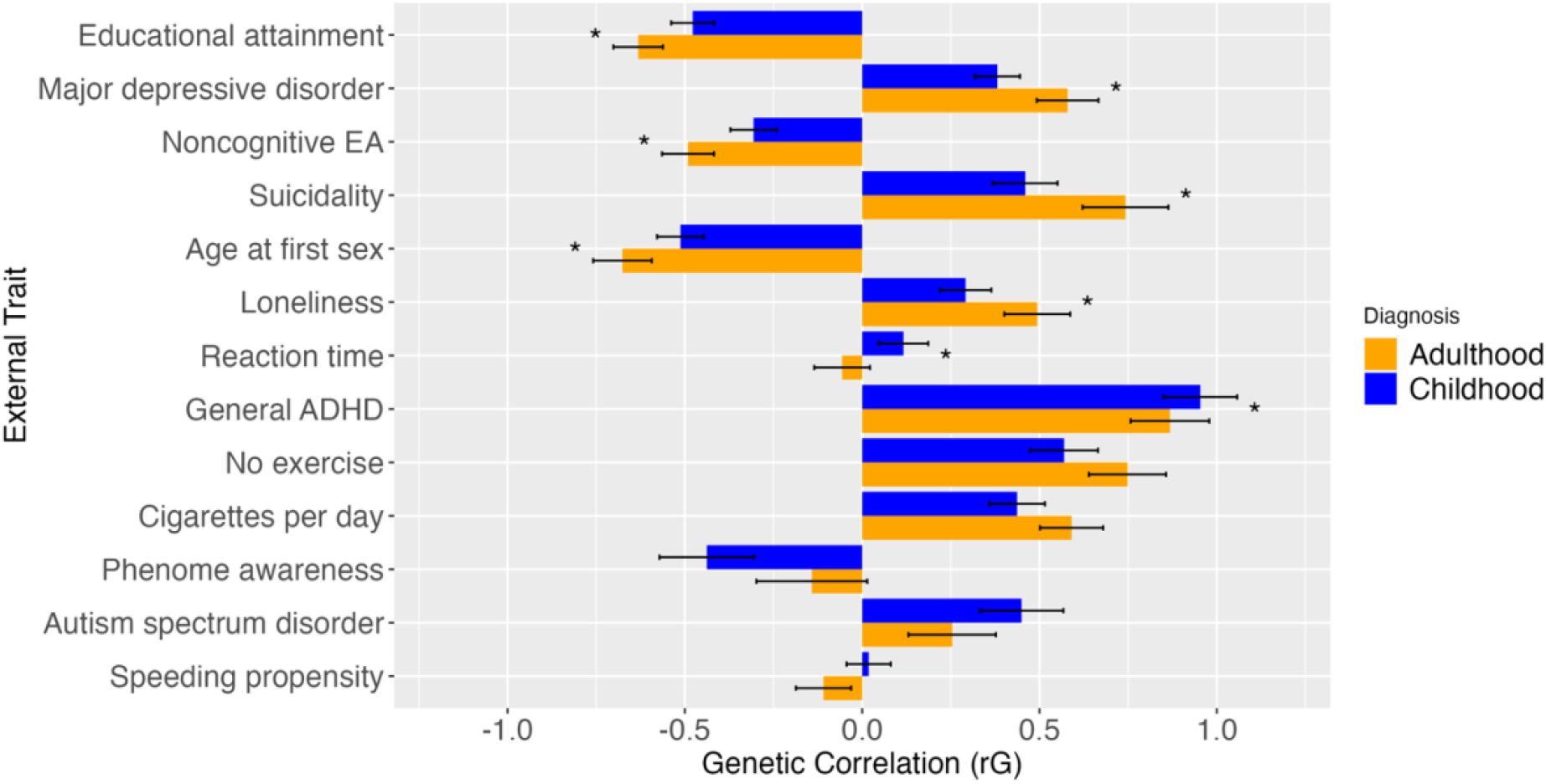
Genetic correlations of adulthood and childhood diagnosed ADHD with external traits. Bonferroni corrected significant differences between adulthood and childhood diagnosed ADHD with the respective external traits are denoted with a star (*) and error bars display 1.96*Standard Error.

Within the cognitive domain, we observed larger negative correlations with educational attainment^23^ for adulthood diagnosed ADHD (*r_g_Adult_* = -0.63 [0.04]) relative to childhood diagnosed (*r_g_Child_*= -0.48 [0.03]; *p_difference_* = 7.12× 10^-9^). Noncognitive skills of EA, reflecting the genetic component of EA that does not overlap with cognitive performance^24^, showed a significantly stronger negative genetic association with adulthood diagnosed ADHD (*r_g_Adult_* = -0.49 [0.04]) relative to childhood diagnosed (*r_g_Child_* = -0.31 [0.03]; *p_difference_* = 2.21× 10^-7^). A childhood diagnosis was also more associated with faster reaction time^25^ (*r_g_Adult_* = -0.06 [0.04] and *r_g_Child_* = 0.12 [0.04], *p_difference_* = 5.72 × 10^-6^). However, we do not observe significant differences for an overall cognitive ability factor (i.e., *g*-factor)^25^ defined by seven cognitive outcomes from UKB.

Within the risk-taking domain, age at first sexual intercourse^26^ showed a stronger negative relationship with adulthood diagnosed ADHD (*r_g_Adult_* = -0.68 [0.04]) than childhood diagnosed ADHD (*r_g_Child_* = -0.51 [0.03]; *p_difference_*= 1.04 × 10^-6^). The psychiatric traits and correlates revealed that MDD^7^ had a stronger positive genetic correlation with adulthood diagnosed ADHD (*r_g_Adult_* = 0.58 [0.04]) relative to childhood diagnosed (*r_g_Child_* = 0.38 [0.03]; *p_difference_*= 6.08 × 10^-8^). Similarly, the genetic signal for suicide attempts^27^ more strongly overlapped with adulthood diagnosed ADHD (*r_g_Adult_* = 0.74 [0.06]) than childhood diagnosed (*r_g_Child_* = 0.46 [0.05]; *p_differenc_*_e_ = 2.88 × 10^-7^). Finally, we highlight that in the interpersonal domain adulthood diagnosed ADHD (*r_g_Adult_* = 0.49 [0.05]) was more strongly associated with loneliness^28^ than childhood diagnosed ADHD (*r_g_Child_* = 0.29 [0.04]; *p_difference_* = 2.55 × 10^-6^). Results for these last three traits (loneliness, suicide attempts, and MDD) can collectively be conceptualized as indexing higher levels of genetic overlap for adult diagnosed ADHD with the internalizing space. Moreover, the genetic signal for MDD, ANX, and PTSD are all strongly overlapping. We went on to leverage the ability of Genomic SEM to model latent, genomic risk factors to examine genetic overlap with an internalizing factor defined by these three disorders.

### Adulthood diagnosed ADHD is More Strongly Associated with Internalizing

The χ^2^ difference test revealed that the significantly larger genetic overlap with adulthood diagnosed ADHD we observed for individual internalizing traits also held for this latent internalizing factor (*r_g_Adult_* = 0.64 [0.05] and *r_g_Child_* = 0.44 [0.04], *p_difference_* = 5.67 × 10^-7^; **Fig. 2**). Although the χ^2^ difference test was not significant for the comparisons including the persistent ADHD phenotype with the child (*p_difference_* = 9.64 × 10^-1^) or adult subgroup (*p_difference_* = 2.41 × 10^-3^), the point estimate was notably more similar to the adult phenotype (*r_g_Persistent_* = 0.65 [0.05]; **Fig. S9)**. We went on to examine whether the significant differences between childhood and adulthood diagnosed ADHD with other external traits is accounted for by the genetic overlap between adulthood diagnosed ADHD and this same internalizing factor. When controlling for shared genetic variance with internalizing, seven traits were found to have significantly different levels of genetic overlap across childhood and adulthood diagnosed ADHD (Table S4). This included five of the traits (e.g., EA) that were identified in the model without internalizing, including an even larger difference between child and adult ADHD with general ADHD. The two new traits that emerged were movement from 10pm – 11pm and ASD,^29^ which evinced stronger association with childhood diagnosed ADHD. Notably, the genetic correlation with ASD and adulthood diagnosed ADHD when controlling for internalizing was near 0 (*r_g_Adult_* = < 0.01 [0.07]; **Fig. 3**). Conversely, the significant differences in genetic correlations with adulthood and childhood diagnosed ADHD in suicidal behavior (*r_g_Adult_* = 0.28 [0.07] and *r_g_Child_* = 0.14 [0.05], *p_difference_* = 3.08 × 10^-3^) and loneliness (*r_g_Adult_* = 0.00 [0.05] and *r_g_Child_* = -0.05 [0.04], *p_difference_* = 9.74 × 10^-2^) became non-significant after accounting for the overlap with the internalizing disorders factor (**Fig. 3**).

**Figure 2.**
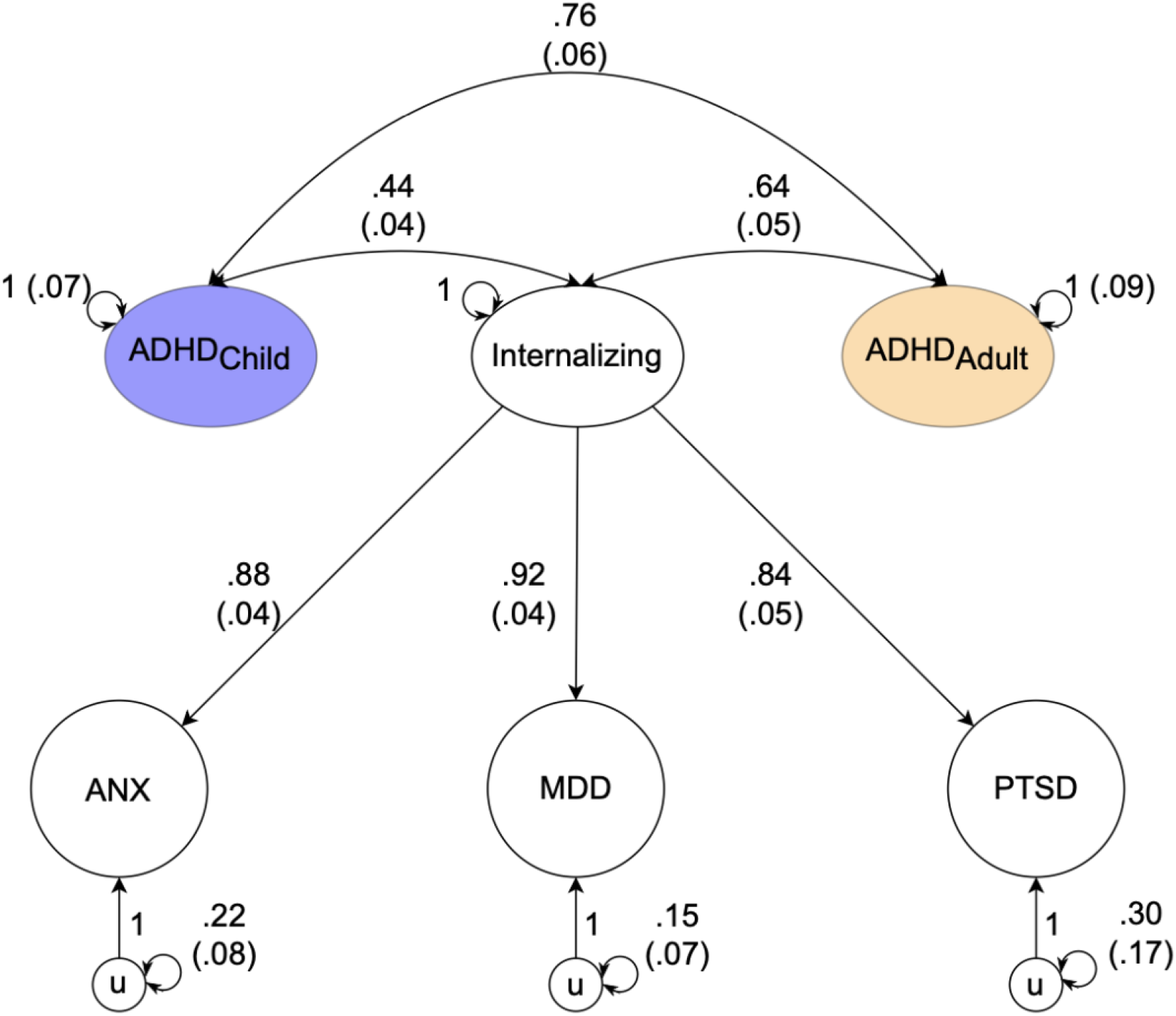
Path diagram of genetic correlations of adulthood and childhood diagnosed ADHD with the internalizing factor. Path diagram of the model used in genomic SEM to confirm that the pattern of larger genetic overlap with adulthood diagnosed ADHD we observe for individual internalizing traits also holds for this latent internalizing factor. In this model internalizing is a common genetic factor of the genetic components of ANX, MDD, and PTSD and u is the residual genetic variance in these phenotypes, not explained by the internalizing factor. Observed variables are represented as squares, and latent variables are represented as circles. The genetic component of each phenotype is represented with a circle as the genetic component is a latent variable that is not directly measured but is inferred using LDSC. Single-headed arrows are regression relations, double-headed arrows connecting back to the same origin are variances, and double-headed arrows connecting two variables are correlations. Paths labeled 1 are fixed to 1.

**Figure 3.**
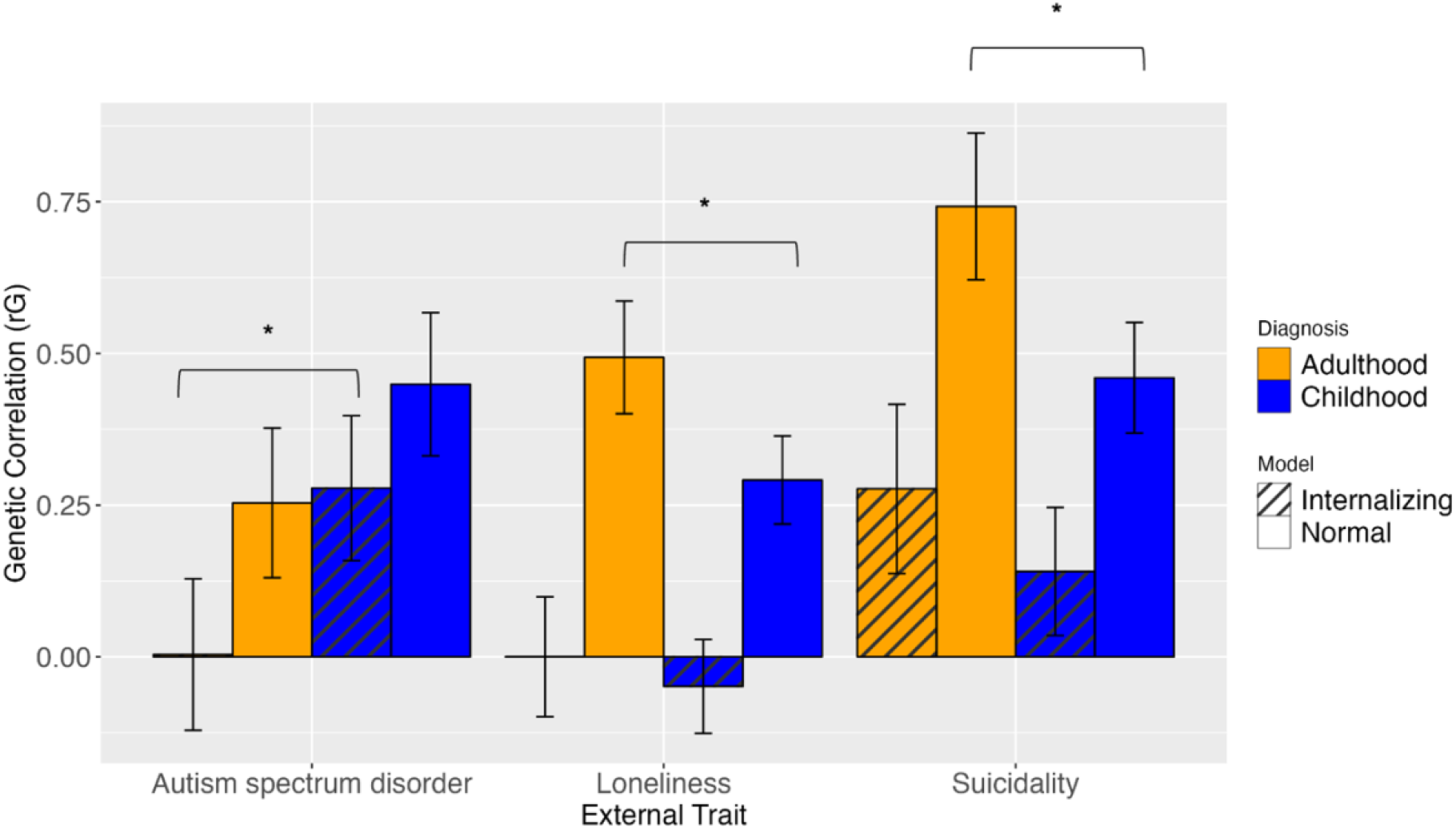
Genetic correlations of adulthood and childhood diagnosed ADHD with external traits. Results without accounting for the overlap of the internalizing factor with ADHD are displayed in the solid bars and results accounting for the overlap of the internalizing factor with ADHD are displayed in the striped bars. Error bars display 1.96*Standard Error.

### Stratified Genomic SEM Identifies Annotations Enriched for Shared Risk

Stratified Genomic SEM identified three annotations that were significantly enriched at the level of a general ADHD factor. (**Fig. 4** and Table S5). This included Fetal Female Brain DNAse (*p* = 1.07 × 10^-7^), Conserved Primate (*p* = 2.01 × 10^-5^), and Fetal Male Brain H3K4me1 (*p* = 6.06 × 10^-5^). We found no significant enrichment for the residual genetic variance unique to ADHD_child_ or ADHD_adult_.

**Figure 4.**
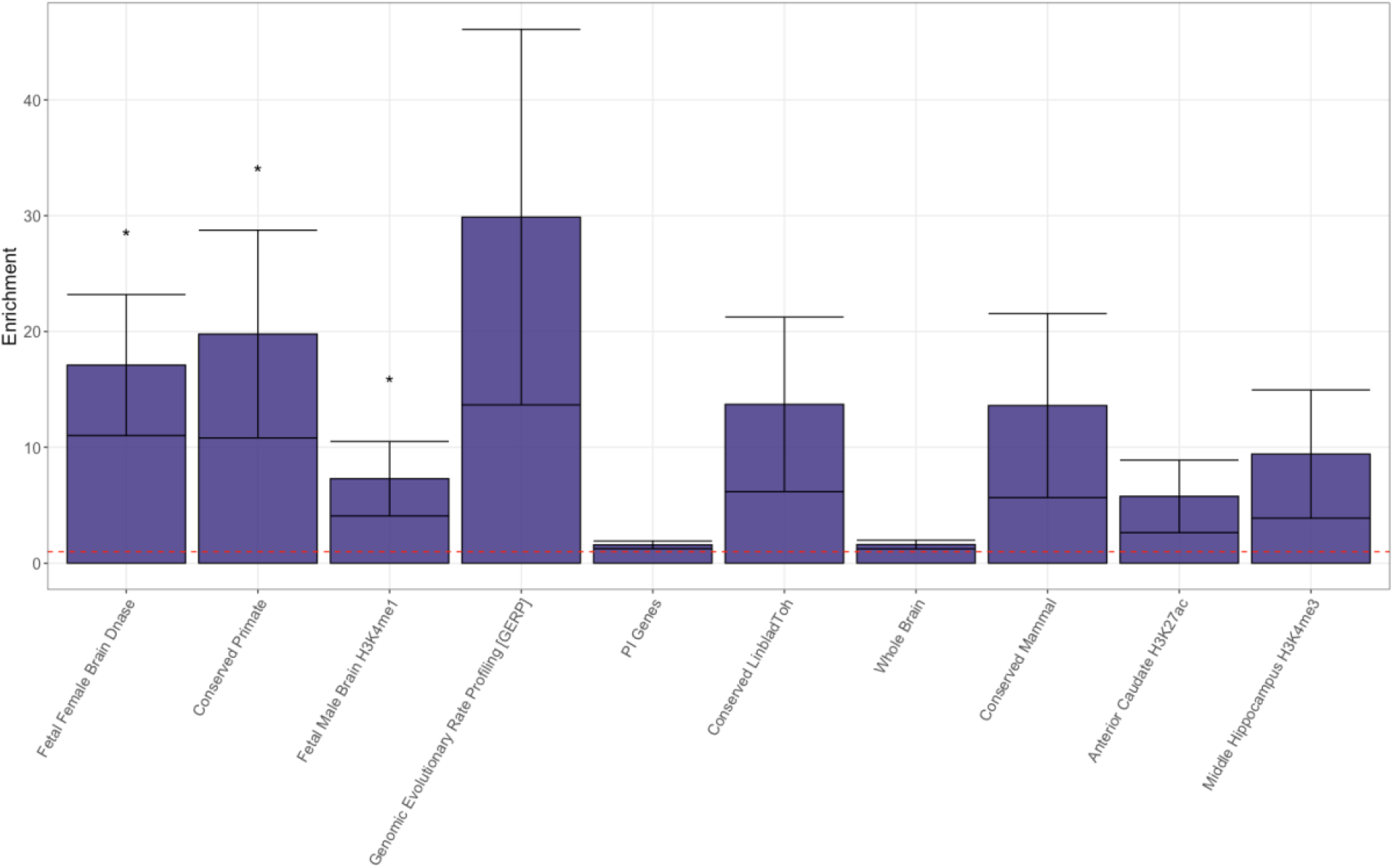
Enrichment of functional annotations in the combined factor of childhood and adulthood diagnosed ADHD. Significant enrichments traits are denoted with a star (*) and error bars display 1.96*Standard Error.

### T-SEM Pinpoints Genes Associated with General ADHD

Univariate TWAS results revealed two genes whose expression was significantly associated with ADHD diagnosed in adulthood and 19 genes for ADHD diagnosed in childhood (Tables S8 and S9). T-SEM then identified 22 unique genes whose expression was associated with a general ADHD factor (**Fig. 5** and Table S11). This included 15 genes that were novel relative to the univariate TWAS of the subgroups and 11 genes that were novel relative to a TWAS of general ADHD that does not distinguish between the different life stages at which ADHD was diagnosed (Table S12). No significant Q_Gene_ hits were identified (Table S10 for top Q_Gene_ results).

**Figure 5.**
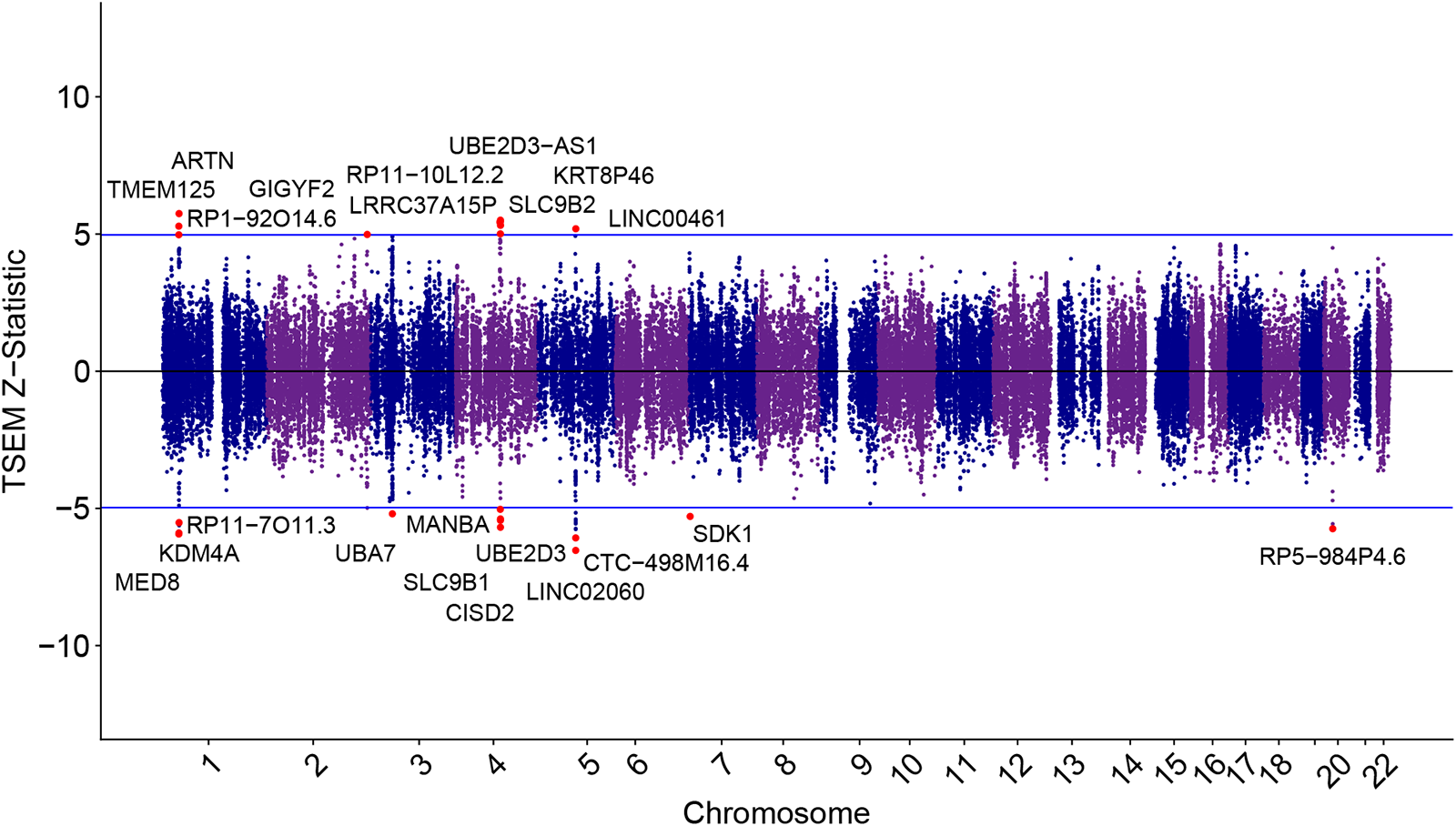
Miami plot for gene expression hits for the combined factor of childhood and adulthood diagnosed ADHD. The upper and lower blue lines represent the Bonferroni corrected significance threshold. Genes surpassing the upper and lower cutoff respectively are upwardly and downwardly regulated respectively in the ADHD factor. The most significant gene across tissue types are labeled and colored as red dots.

## DISCUSSION

The current study revealed a mixture of convergent and divergent genetic signal in ADHD diagnosed in adulthood and childhood. At the level of genetic overlap with external correlates the primary differences were identified for cognitive and internalizing outcomes. Within the cognitive domain, ADHD diagnosed in adulthood showed a stronger negative association with educational attainment and non-cognitive skills of educational attainment. This divergent relationship did not hold for a range of cognitive traits, including for a *g-*factor of general intelligence. The collective pattern of findings then indicates that adult diagnosed ADHD genetically overlaps specifically with the noncognitive aspects that lead to success in school settings.

Within the internalizing space, we show that adulthood diagnosed ADHD has a sizeable and more positive association with MDD, suicidal behavior, loneliness, and a latent internalizing factor defined by MDD, PTSD, and ANX. One possible interpretation of this pattern of findings is that adult diagnosed ADHD can, in some instances, reflect a misdiagnosis that occurs at higher rates than in children. This is supported by the fact that the internalizing space shares overlapping symptoms, including difficulty concentrating and restlessness. An alternative interpretation is that ADHD diagnosed in adulthood is not a misdiagnosis, but rather is more comorbid with other internalizing disorders. For example, it could be that ADHD diagnosed in adulthood is more disruptive to daily living or the absence of a diagnosis until that later life stage increases risk for other disorders. In support of this, adults with ADHD often display high rates of general mental health symptoms, such as anxiety and depression.^30^ However, we observe such high levels of genetic overlap despite the fact that the genetic overlap is estimated from separate samples of individuals with internalizing disorders. Thus, some causal link between ADHD and internalizing is unlikely to explain the bulk of our findings as this would require ADHD to be one of the primary risk factors for internalizing (or vice versa) to be reflected in estimates of genetic overlap from independent participant samples.

Follow-up models accounting for the shared variance with an internalizing factor provided two additional sets of findings for interpreting the emergent etiological picture. First, we find that the internalizing factor explains the larger genetic overlap between adult diagnosed ADHD and two clinical correlates of internalizing: loneliness and suicidal behavior. Second, childhood diagnosed ADHD showed a much stronger association with both ADHD and ASD than adult diagnosed ADHD when controlling for shared variance with internalizing. Moreover, the genetic correlation with adult diagnosed and ASD was estimated near 0. This indicates that the neurodevelopmental signal captured in the adult diagnosed ADHD GWAS is greatly diminished, or entirely absent, when removing shared genetic variance with internalizing, further supporting a hypothesis of increased levels of misdiagnosis in adulthood.

Evidence against the misdiagnosis interpretation came from follow-up models examining the correlation with persistent ADHD and internalizing. More specifically, we observe that the point estimate for the genetic correlation with internalizing was more similar for adulthood and persistent ADHD as compared to childhood ADHD. Although the difference between persistent and childhood ADHD was not significant, we were generally underpowered for analyses using the persistent phenotype. As persistent ADHD is arguably less likely to reflect misdiagnosis given its original diagnosis in childhood these tentative results indicate that internalizing disorders may play a significant role in fostering the persistence of ADHD into adulthood. However, it is important to note that these findings present an alternative perspective rather than definitively pointing to one mechanism over the other and should be re-evaluated once power is sufficient to gain a more comprehensive understanding. Another possible mechanism for this internalizing overlap with adulthood diagnosed ADHD are sex differences in symptom patterns as well as age of diagnosis. Notably, 41% of the adulthood diagnosed cases were female, in contrast to 23% of the childhood diagnosed cases^5^. At the same time, females with ADHD are more likely to exhibit internalizing symptoms which are also more characteristic for older individuals with ADHD, while males are more likely to exhibit externalizing symptoms, more common among younger individuals with ADHD^31^. This interplay could inflate the association of adulthood diagnosed ADHD with internalizing disorders due to a cooccurrence of delayed diagnosis and increased internalizing symptoms among females. Future research should assess the connection between sex differences in ADHD and age of diagnosis to explore alternative explanations for these patterns beyond misdiagnosis. Irrespective of whether a misdiagnosis or a clinically relevant distinction between ADHD diagnosed in childhood and adulthood is the cause for their divergence, etiological differences between the two ages of onset might inform the way we treat individuals with ADHD.

Stratified Genomic SEM revealed three annotations that were significantly enriched for the shared pathways between childhood and adulthood diagnosed ADHD: fetal female brain DNAse, fetal male brain H3K4me1, and conserved primate. These fetal annotations^32^ and conserved regions^11^ have been identified for human complex traits more generally. We, therefore, refrain from interpreting these annotations as evidence supporting or disproving the grouping of child and adult ADHD within a single diagnostic class.

T-SEM analyses identified 22 genes significantly associated with shared ADHD risk. Highlighting the ability of multivariate approaches to yield novel discoveries, this included 15 novel genes relative to univariate TWAS of child or adult diagnosed ADHD and 11 novel genes relative to a TWAS of general ADHD. The five most significant genes have been previously linked to ADHD and related phenotypes. *LINC02060* and *CTC-498M16.4* have been found to be related to ADHD and *CTC-498M16.4* has been linked to additional phenotypes, including depression and sleep characteristics^33, 34^. *MED8* has been found to bind to regulatory elements and is a gene of interest for both ADHD and schizophrenia susceptibility^35, 36^. *KDM4A* is related to disruptive behavior disorders in the context of ADHD^37^. Finally, *ARTN* supports the survival, development, methylation, and differentiation of neurons and has been found to be related to ADHD and schizophrenia^33, 36, 38^. Identifying these genes in the contexts of these analyses clarifies their role as influencing shared pathways across these two ADHD subgroups. Likely due to limited power, no genes were significant for the Q_Gene_ heterogeneity statistic applied here to identify genes with divergent patterns of association with childhood and adulthood diagnosed ADHD. *DNM1* and *CRIM1* displayed some of the strongest patterns of divergence, with stronger associations with adulthood diagnosed ADHD and childhood diagnosed ADHD respectively. *DNM1* has been associated with developmental delay and epilepsy^39^ and *CRIM1* has been identified to have a role in development as well as motor neuron differentiation and survival^40^.

### Limitations

All summary statistics in this study are limited to European ancestry because sample sizes are lacking for other ancestries. However, ADHD is a global problem^41^ and in order to understand this disorder and its relationship with other disorders, traits, and behaviors and to treat individuals from around the world, we need to expand our sample sizes in underrepresented populations and include them in our research.

The absence of enrichment of the residual variance of the ADHD subgroups in the Stratified Genomic SEM analyses and Q_Gene_ hits in T-SEM analyses likely reflects reduced power relative to the genome-wide analyses. These results should be re-evaluated as the ADHD GWAS sample sizes increase. Finally, since our analyses are based on genetic correlations, any limitations applying to genetic correlations apply here as well. For example, it has recently been highlighted that genetic correlations can be biased upward by cross-trait assortative mating^42^. Cross-trait assortative mating is the mechanism in which individuals who score higher than average on one trait mate with others who score higher than average on a different trait. This limitation requires special attention since these biases do not have the same magnitude between any pair of included traits. However, because the genetic correlations of many traits included, especially psychiatric traits, are substantial, it is highly unlikely that the pattern of findings here is entirely due to cross-trait assortative mating.^43^

### Conclusions

Although effective treatment approaches to ADHD are available, strategies for finding the right treatment are often based on a trial-and-error approach. This can lead patients to give up before finding the appropriate intervention^44^. Disentangling pathways, symptoms and comorbidities specific to different subtypes and clinical presentations of ADHD, such as childhood and adulthood diagnosed subtypes, might help to improve treatment outcomes by more quickly identifying medications and therapies that are likely effective for specific groups of individuals. The current findings indicate that ADHD diagnosed in adulthood is far more genetically similar to internalizing disorders and clinical correlates as compared to childhood diagnosed ADHD. Whether this reflects pervasive misdiagnosis or distinct patterns of genetic risk across these two groups, identifying these differences highlights the clinical and etiological importance of distinguishing these subgroups within this overarching disorder class.

## Data Availability

The summary statistics for child and adult diagnosed ADHD are available from the iPSYCH website. The source for each external trait is listed in Supplementary Table 1.

https://ipsych.dk/en/research/downloads

## Acknowledgements

SB, ADG, and JML are supported by NIMH Grant R01MH120219. ADG and IFF are supported by NIA Grant RF1AG073593.

## REFERENCES

1. Polanczyk G, de Lima MS, Horta BL, Biederman J, Rohde LA. The Worldwide Prevalence of ADHD: A Systematic Review and Metaregression Analysis. Am J Psychiatry. Published online 2007.

2. Baselmans BML, Yengo L, Van Rheenen W, Wray NR. Risk in Relatives, Heritability, SNP-Based Heritability, and Genetic Correlations in Psychiatric Disorders: A Review. Biological Psychiatry. 2021;89(1):11–19. doi:10.1016/j.biopsych.2020.05.034

3. Miller CJ, Newcorn JH, Halperin JM. Fading Memories: Retrospective Recall Inaccuracies in ADHD. J Atten Disord. 2010;14(1):7–14. doi:10.1177/1087054709347189

4. Agnew-Blais JC, Belsky DW, Caspi A, et al. Polygenic Risk and the Course of Attention-Deficit/Hyperactivity Disorder From Childhood to Young Adulthood: Findings From a Nationally Representative Cohort. Journal of the American Academy of Child & Adolescent Psychiatry. 2021;60(9):1147–1156. doi:10.1016/j.jaac.2020.12.033

5. Rajagopal VM, Duan J, Vilar-Ribó L, et al. Differences in the genetic architecture of common and rare variants in childhood, persistent and late-diagnosed attention-deficit hyperactivity disorder. Nat Genet. 2022;54(8):1117–1124. doi:10.1038/s41588-022-01143-7

6. Bulik-Sullivan B, Finucane HK, Anttila V, et al. An atlas of genetic correlations across human diseases and traits. Nat Genet. 2015;47(11):1236–1241. doi:10.1038/ng.3406

7. Howard DM, Adams MJ, Clarke TK, et al. Genome-wide meta-analysis of depression identifies 102 independent variants and highlights the importance of the prefrontal brain regions. Published online 2019:33.

8. Purves KL, Coleman JRI, Meier SM, et al. A major role for common genetic variation in anxiety disorders. Mol Psychiatry. 2020;25(12):3292–3303. doi:10.1038/s41380-019-0559-1

9. Nievergelt CM, Maihofer AX, Klengel T, et al. International meta-analysis of PTSD genome-wide association studies identifies sex- and ancestry-specific genetic risk loci. Nat Commun. 2019;10(1):4558. doi:10.1038/s41467-019-12576-w

10. Grotzinger AD, Mallard TT, Akingbuwa WA, et al. Genetic Architecture of 11 Major Psychiatric Disorders at Biobehavioral, Functional Genomic, and Molecular Genetic Levels of Analysis. Genetic and Genomic Medicine; 2020. doi:10.1101/2020.09.22.20196089

11. ReproGen Consortium, Schizophrenia Working Group of the Psychiatric Genomics Consortium, The RACI Consortium, et al. Partitioning heritability by functional annotation using genome-wide association summary statistics. Nat Genet. 2015;47(11):1228–1235. doi:10.1038/ng.3404

12. Hujoel MLA, Gazal S, Hormozdiari F, Van De Geijn B, Price AL. Disease Heritability Enrichment of Regulatory Elements Is Concentrated in Elements with Ancient Sequence Age and Conserved Function across Species. The American Journal of Human Genetics. 2019;104(4):611–624. doi:10.1016/j.ajhg.2019.02.008

13. The GTEx Consortium, Ardlie KG, Deluca DS, et al. The Genotype-Tissue Expression (GTEx) pilot analysis: Multitissue gene regulation in humans. Science. 2015;348(6235):648–660. doi:10.1126/science.1262110

14. Pers TH, Karjalainen JM, Chan Y, et al. Biological interpretation of genome-wide association studies using predicted gene functions. Nat Commun. 2015;6(1):5890. doi:10.1038/ncomms6890

15. Karczewski KJ, Francioli LC, Tiao G, et al. The mutational constraint spectrum quantified from variation in 141,456 humans. Nature. 2020;581(7809):434–443. doi:10.1038/s41586-020-2308-7

16. Roadmap Epigenomics Consortium, Kundaje A, Meuleman W, et al. Integrative analysis of 111 reference human epigenomes. Nature. 2015;518(7539):317–330. doi:10.1038/nature14248

17. Grotzinger AD, de la Fuente J, Davies G, Nivard MG, Tucker-Drob EM. Transcriptome-wide and stratified genomic structural equation modeling identify neurobiological pathways shared across diverse cognitive traits. Nat Commun. 2022;13(1):6280. doi:10.1038/s41467-022-33724-9

18. Gusev A, Ko A, Shi H, et al. Integrative approaches for large-scale transcriptome-wide association studies. Nat Genet. 2016;48(3):245–252. doi:10.1038/ng.3506

19. Lonsdale J, Thomas J, Salvatore M, et al. The Genotype-Tissue Expression (GTEx) project. Nat Genet. 2013;45(6):580–585. doi:10.1038/ng.2653

20. Hoffman GE, Bendl J, Voloudakis G, et al. CommonMind Consortium provides transcriptomic and epigenomic data for Schizophrenia and Bipolar Disorder. Sci Data. 2019;6(1):180. doi:10.1038/s41597-019-0183-6

21. Gandal MJ, Zhang P, Hadjimichael E, et al. Transcriptome-wide isoform-level dysregulation in ASD, schizophrenia, and bipolar disorder. Science. 2018;362(6420):eaat8127. doi:10.1126/science.aat8127

22. ADHD Working Group of the Psychiatric Genomics Consortium (PGC), Early Lifecourse & Genetic Epidemiology (EAGLE) Consortium, 23andMe Research Team, et al. Discovery of the first genome-wide significant risk loci for attention deficit/hyperactivity disorder. Nat Genet. 2019;51(1):63–75. doi:10.1038/s41588-018-0269-7

23. Okbay A, Wu Y, Wang N, et al. Polygenic prediction of educational attainment within and between families from genome-wide association analyses in 3 million individuals. Nat Genet. Published online March 31, 2022. doi:10.1038/s41588-022-01016-z

24. Demange PA, Malanchini M, Mallard TT, et al. Investigating the genetic architecture of noncognitive skills using GWAS-by-subtraction. Nat Genet. 2021;53(1):35–44. doi:10.1038/s41588-020-00754-2

25. de la Fuente J, Davies G, Grotzinger AD, Tucker-Drob EM, Deary IJ. A general dimension of genetic sharing across diverse cognitive traits inferred from molecular data. Nat Hum Behav. 2021;5(1):49–58. doi:10.1038/s41562-020-00936-2

26. Watanabe K, Stringer S, Frei O, et al. A global overview of pleiotropy and genetic architecture in complex traits. Nat Genet. 2019;51(9):1339–1348. doi:10.1038/s41588-019-0481-0

27. Mullins N, Kang J, Campos AI, et al. Dissecting the Shared Genetic Architecture of Suicide Attempt, Psychiatric Disorders, and Known Risk Factors. Biological Psychiatry. 2022;91(3):313–327. doi:10.1016/j.biopsych.2021.05.029

28. Day FR, Ong KK, Perry JRB. Elucidating the genetic basis of social interaction and isolation. Nat Commun. 2018;9(1):2457. doi:10.1038/s41467-018-04930-1

29. Autism Spectrum Disorder Working Group of the Psychiatric Genomics Consortium, BUPGEN, Major Depressive Disorder Working Group of the Psychiatric Genomics Consortium, et al. Identification of common genetic risk variants for autism spectrum disorder. Nat Genet. 2019;51(3):431–444. doi:10.1038/s41588-019-0344-8

30. Skirrow C, Asherson P. Emotional lability, comorbidity and impairment in adults with attention-deficit hyperactivity disorder. Journal of Affective Disorders. 2013;147(1-3):80–86. doi:10.1016/j.jad.2012.10.011

31. Franke B, Michelini G, Asherson P, et al. Live fast, die young? A review on the developmental trajectories of ADHD across the lifespan. European Neuropsychopharmacology. 2018;28(10):1059–1088. doi:10.1016/j.euroneuro.2018.08.001

32. Finucane HK, Reshef YA, Anttila V, et al. Heritability enrichment of specifically expressed genes identifies disease-relevant tissues and cell types. Nat Genet. 2018;50(4):621–629. doi:10.1038/s41588-018-0081-4

33. Liao C, Laporte AD, Spiegelman D, et al. Transcriptome-wide association study of attention deficit hyperactivity disorder identifies associated genes and phenotypes. Nat Commun. 2019;10(1):4450. doi:10.1038/s41467-019-12450-9

34. O’Connell KS, Frei O, Bahrami S, et al. Characterizing the Genetic Overlap Between Psychiatric Disorders and Sleep-Related Phenotypes. Biological Psychiatry. 2021;90(9):621–631. doi:10.1016/j.biopsych.2021.07.007

35. Alonso-Gonzalez A, Calaza M, Rodriguez-Fontenla C, Carracedo A. Gene-based analysis of ADHD using PASCAL: a biological insight into the novel associated genes. BMC Med Genomics. 2019;12(1):143. doi:10.1186/s12920-019-0593-5

36. Reay WR, Cairns MJ. Pairwise common variant meta-analyses of schizophrenia with other psychiatric disorders reveals shared and distinct gene and gene-set associations. Transl Psychiatry. 2020;10(1):134. doi:10.1038/s41398-020-0817-7

37. Demontis D, Walters RK, Rajagopal VM, et al. Risk variants and polygenic architecture of disruptive behavior disorders in the context of attention-deficit/hyperactivity disorder. Nat Commun. 2021;12(1):576. doi:10.1038/s41467-020-20443-2

38. Ilieva M, Nielsen J, Korshunova I, et al. Artemin and an Artemin-Derived Peptide, Artefin, Induce Neuronal Survival, and Differentiation Through Ret and NCAM. Front Mol Neurosci. 2019;12:47. doi:10.3389/fnmol.2019.00047

39. Michetti C, Falace A, Benfenati F, Fassio A. Synaptic genes and neurodevelopmental disorders: From molecular mechanisms to developmental strategies of behavioral testing. Neurobiology of Disease. 2022;173:105856. doi:10.1016/j.nbd.2022.105856

40. Kolle G, Georgas K, Holmes GP, Little MH, Yamada T. CRIM1, a novel gene encoding a cysteine-rich repeat protein, is developmentally regulated and implicated in vertebrate CNS development and organogenesis. Mechanisms of Development. 2000;90(2):181–193. doi:10.1016/S0925-4773(99)00248-8

41. GBD 2019 Mental Disorders Collaborators. Global, regional, and national burden of 12 mental disorders in 204 countries and territories, 1990–2019: a systematic analysis for the Global Burden of Disease Study 2019. The Lancet Psychiatry. 2022;9(2):137–150. doi:10.1016/S2215-0366(21)00395-3

42. Border R, Athanasiadis G, Buil A, et al. Cross-trait assortative mating is widespread and inflates genetic correlation estimates. Published online 2022.

43. Grotzinger AD, Keller MC. Potential bias in genetic correlations. Science. 2022;378(6621):709–710. doi:10.1126/science.ade8002

44. Mamiya P, Arnett A, Stein M. Precision Medicine Care in ADHD: The Case for Neural Excitation and Inhibition. Brain Sciences. 2021;11(1):91. doi:10.3390/brainsci11010091

