## Supplementary material for "Examining Differences in the Genetic and Functional Architecture of ADHD Diagnosed in Childhood and Adulthood": Online Supplement

**Supplementary Note**

***Dataset Quality Control.*** Only GWAS summary statistics for traits that had a $h_{SNP}^{2}$ *Z*-statistic > 4 were included in accordance with the suggested cutoffs provided by the original LDSC developers for producing interpretable estimates of genetic overlap across pairs of traits ^1^. The cognitive traits battery is the exception to this with two traits that did not reach the *Z*-statistic threshold of 4 being included in a more well-powered *g*-factor model that followed the common factor specification from^2^. Prior to running *ldsc,* all GWAS summary statistics were subjected to a standardized quality control pipeline using the *munge* function. This function works to restrict to HapMap3 SNPs, align all GWAS summary statistics relative to the same reference allele, and to filter to a minor allele frequency (MAF) > 1% and imputation quality (INFO score) > 0.9 when this information was available. Multivariable *ldsc* was run using LD-scores that were calculated using the European subsample of the 1000 Genomes Phase 3 project excluding the MHC region.

For binary traits (e.g., the ADHD subgroups), estimates were converted to the more interpretable liability scale that accounts for both the continuous (liability) distribution of genetic risk and sample ascertainment. This liability scale conversion was calculated using the population prevalence from the paper for the corresponding GWAS. If the population prevalence was not given in the original paper, a population prevalence reflective of the participant sample was used. In addition, we input the sum of effective sample size across the cohorts contributing as we have recently shown this to produce more accurate estimates of liability-scale heritability^3^. We then entered a sample prevalence of .5 to reflect the fact that sample ascertainment was already corrected for by calculating the sum of effective sample size. For Autism Spectrum Disorder, effective sample size was calculated directly from the GWAS data^3^ as the effective sample size implied by trio data is smaller than the observed sum of effective sample size. This is due to the reduced power due to having parents as controls that have a higher polygenic load for the phenotype than standard control subjects ^4^.

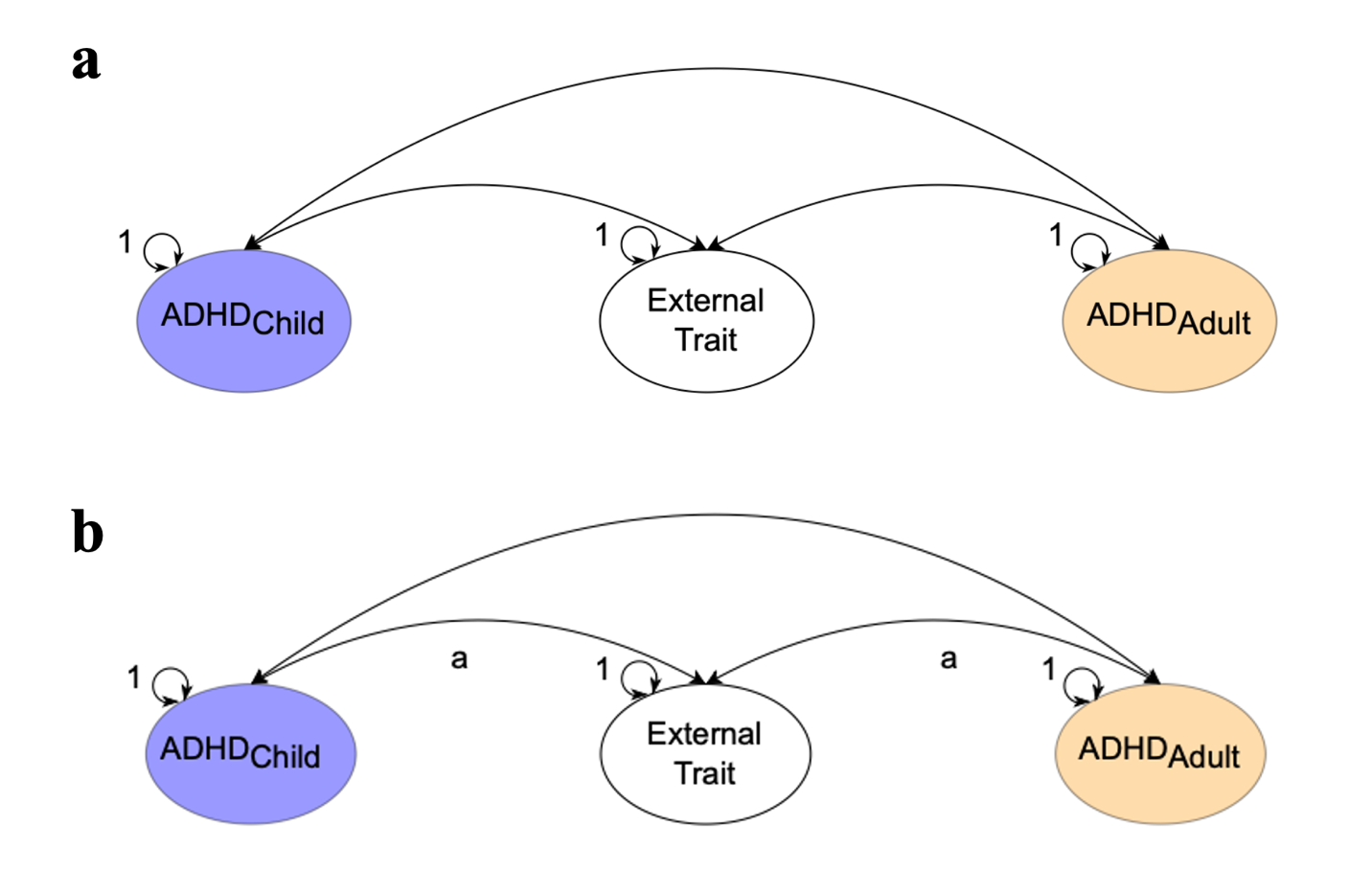

**Supplementary Figure 1. Model Comparisons for Genetic Correlations of Childhood and Adulthood Diagnosed ADHD with External Traits. a**, unconstrained model **b**, model constraining the genetic correlations of adulthood and childhood diagnosed ADHD with the external traits to be equal. The genetic components of ADHD and external traits are inferred variables that are represented as ovals. Correlations between variables are represented as two-headed arrows linking the variables. (Residual) variances of a variable are represented as a two-headed arrow connecting the variable to itself. Paths labeled 1 are fixed to 1 and paths labeled with the same letter are fixed to be equal.

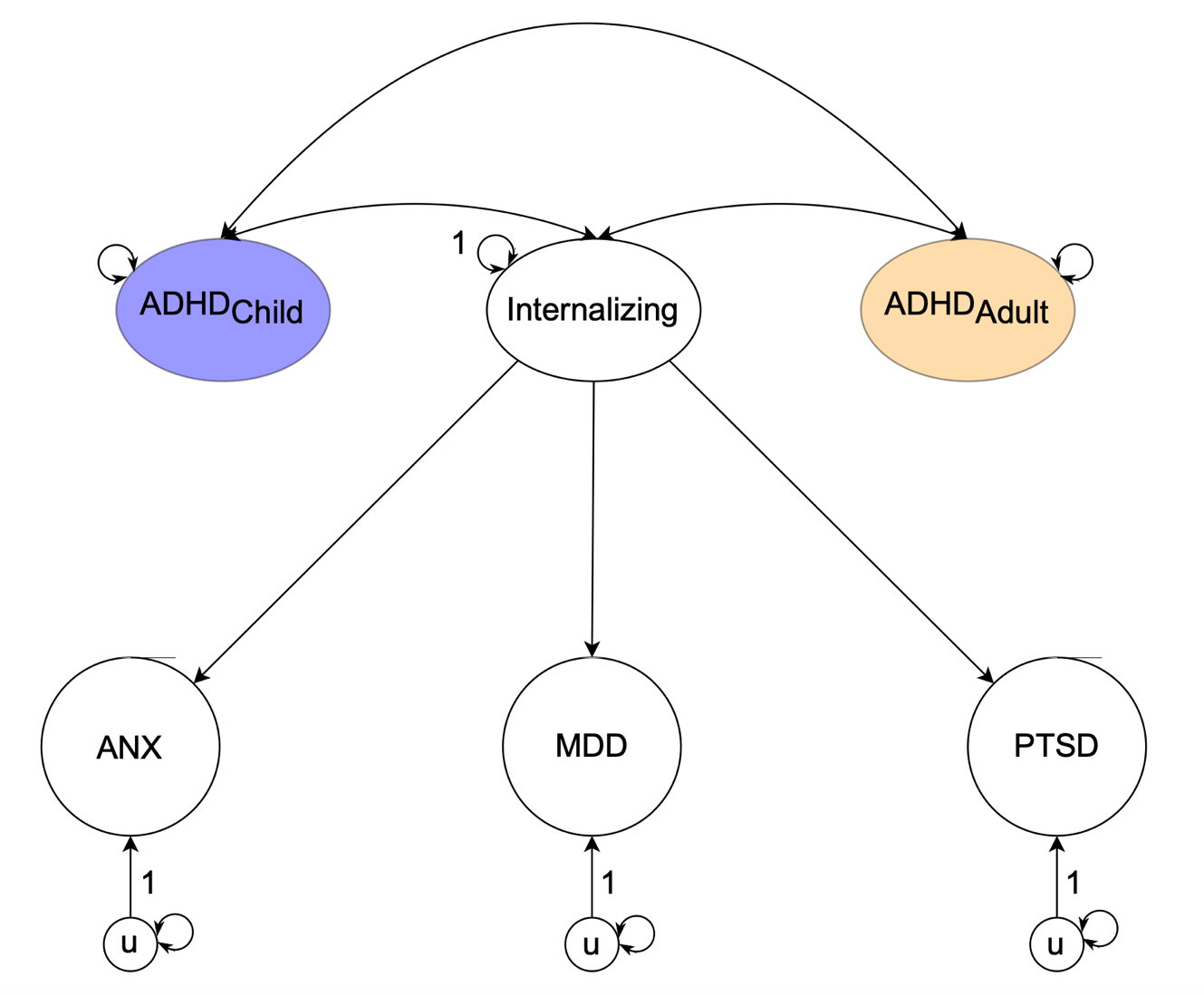

**Supplementary Figure 2. Genetic Correlations of Childhood and Adulthood Diagnosed ADHD with an Internalizing Factor.** Model for deriving the genetic correlations of childhood and adulthood diagnosed ADHD with an internalizing factor, composed of ANX, MDD, and PTSD. The genetic components of disorders and factors are inferred variables that are represented as ovals. Correlations between variables are represented as two-headed arrows linking the variables. (Residual) variances of a variable are represented as a two-headed arrow connecting the variable to itself.

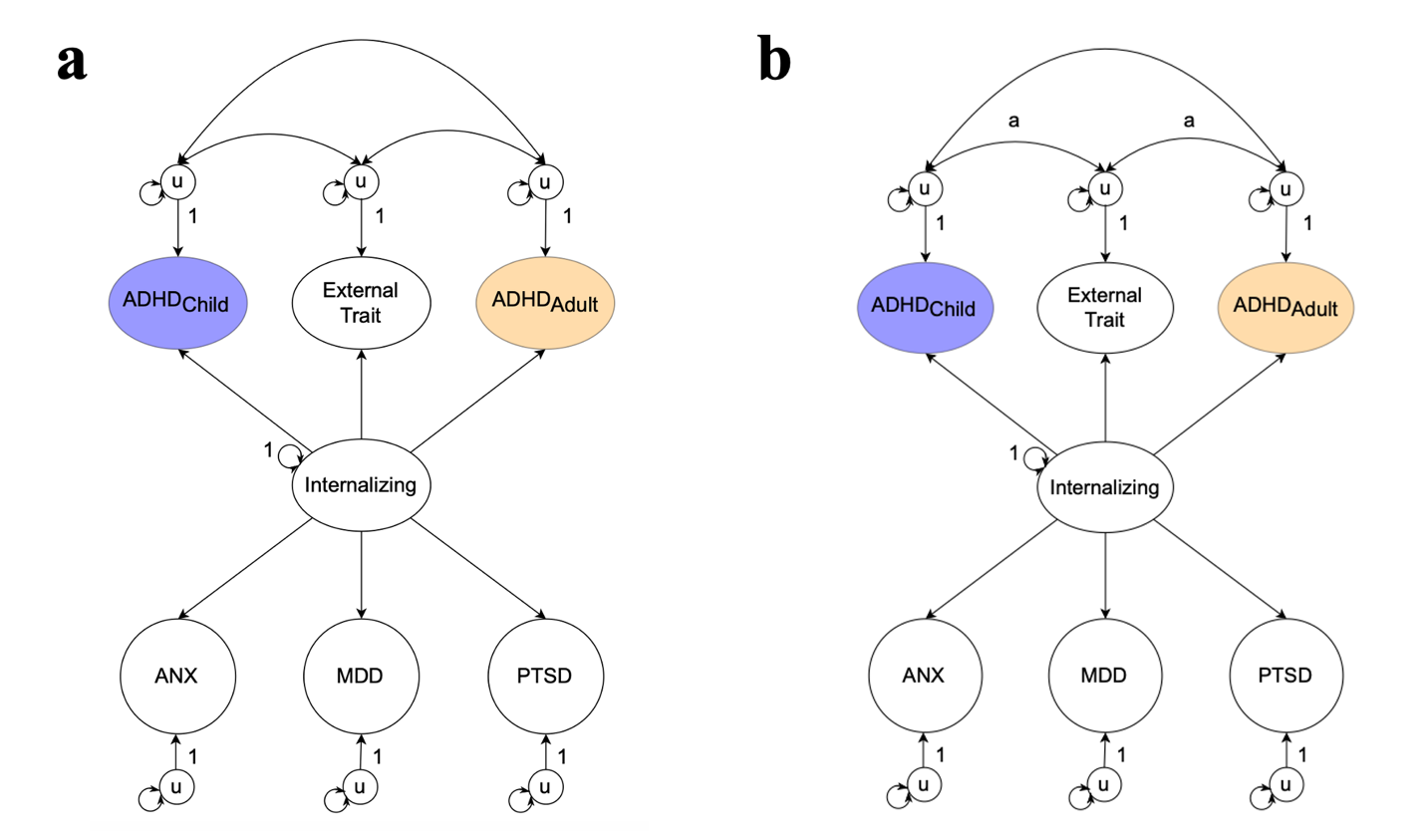

**Supplementary Figure 3. Model Comparisons for Genetic Correlations of Childhood and Adulthood Diagnosed ADHD with External Traits Accounting for Overlap with the Internalizing Factor.** **a**, unconstrained model **b**, model constraining the genetic correlations of adulthood and childhood diagnosed ADHD with the external traits to be equal. The genetic components of the disorders, the internalizing factor, and the external traits are inferred variables that are represented as ovals. Correlations between variables are represented as two-headed arrows linking the variables. (Residual) variances of a variable are represented as a two-headed arrow connecting the variable to itself. Paths labeled 1 are fixed to 1 and paths labeled with the same letter are fixed to be equal.

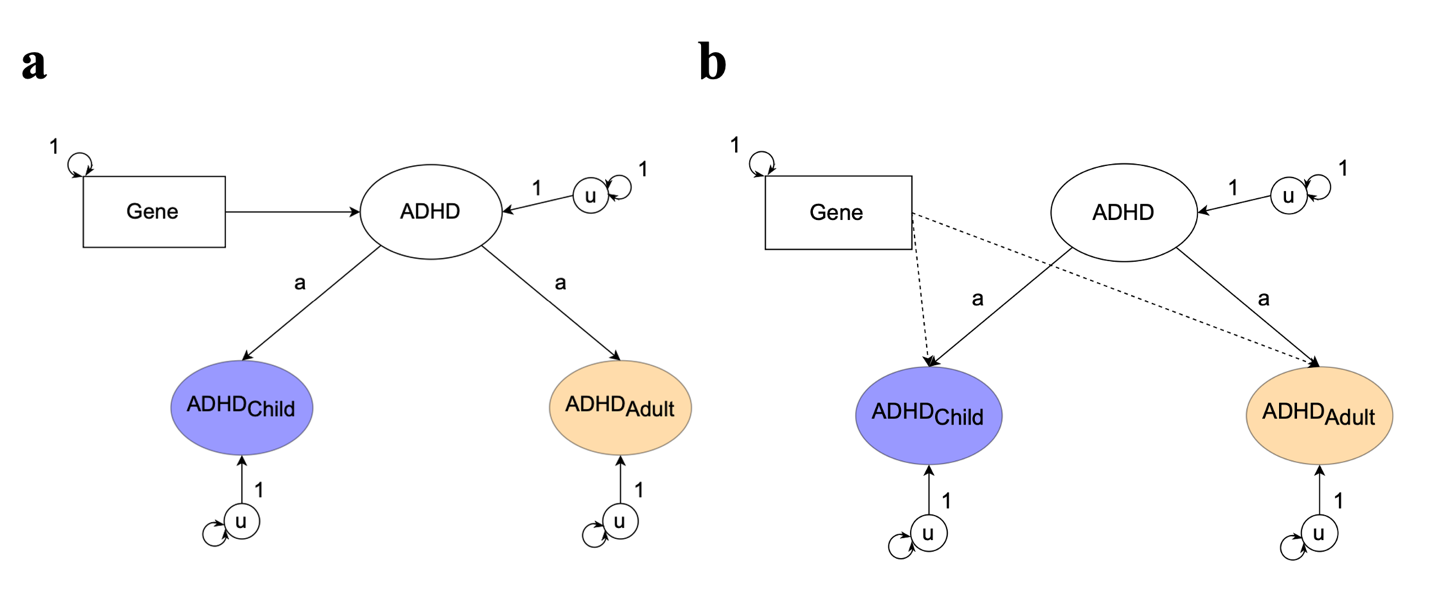
**Supplementary Figure 4. Model comparison for the QGene TSEM-Analyses. a**, Common pathway model where genes operate through the combine ADHD inferring the gene is associated with a shared risk between ADHD diagnosed in childhood and adulthood **b**, Independent pathways model where genes operate on ADHD diagnosed in adulthood and childhood individually, inferring that the gene’s effects are specific to either age of diagnosis. The genetic component of childhood and adulthood diagnosed ADHD, as well as the combined ADHD factor are represented with an oval as the genetic component is a latent variable that is inferred. SNPs are directly measured and are therefore represented as squares. Single-headed arrows are regression relations, and (residual) variances of a variable are represented as a two-headed arrow connecting the variable to itself. Paths labeled 1 are fixed to 1. All unlabeled paths represent freely estimated model parameters.

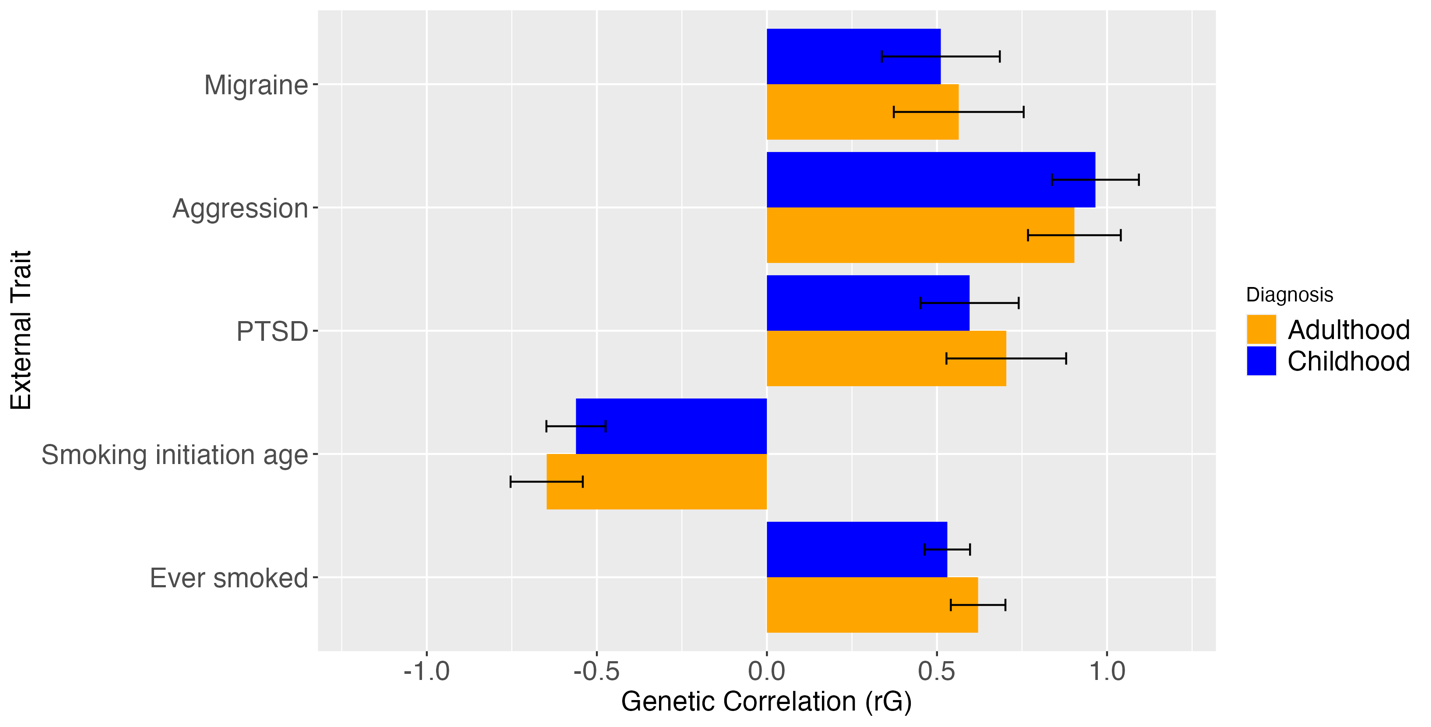

**Supplementary Figure 5. Genetic correlations of adulthood and childhood diagnosed ADHD with external traits.** High genetic correlations (above .5) that show differences between genetic correlations with childhood and adulthood diagnosed ADHD of less than .15. Error bars display 1.96*Standard Error.

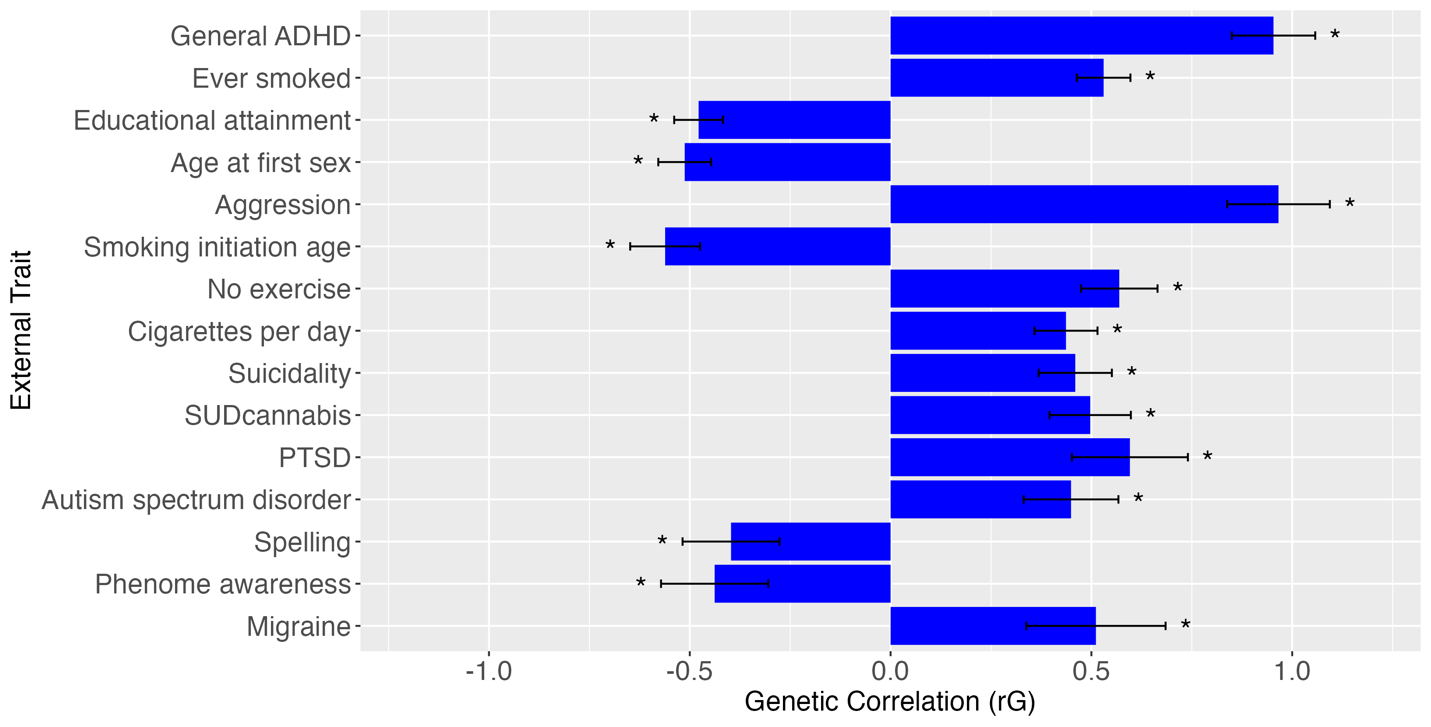

**Supplementary Figure 6. 15 most significant genetic correlations with childhood diagnosed ADHD.** Sorted by smallest to largest p-value from top to bottom. Error bars display 1.96*Standard Error.

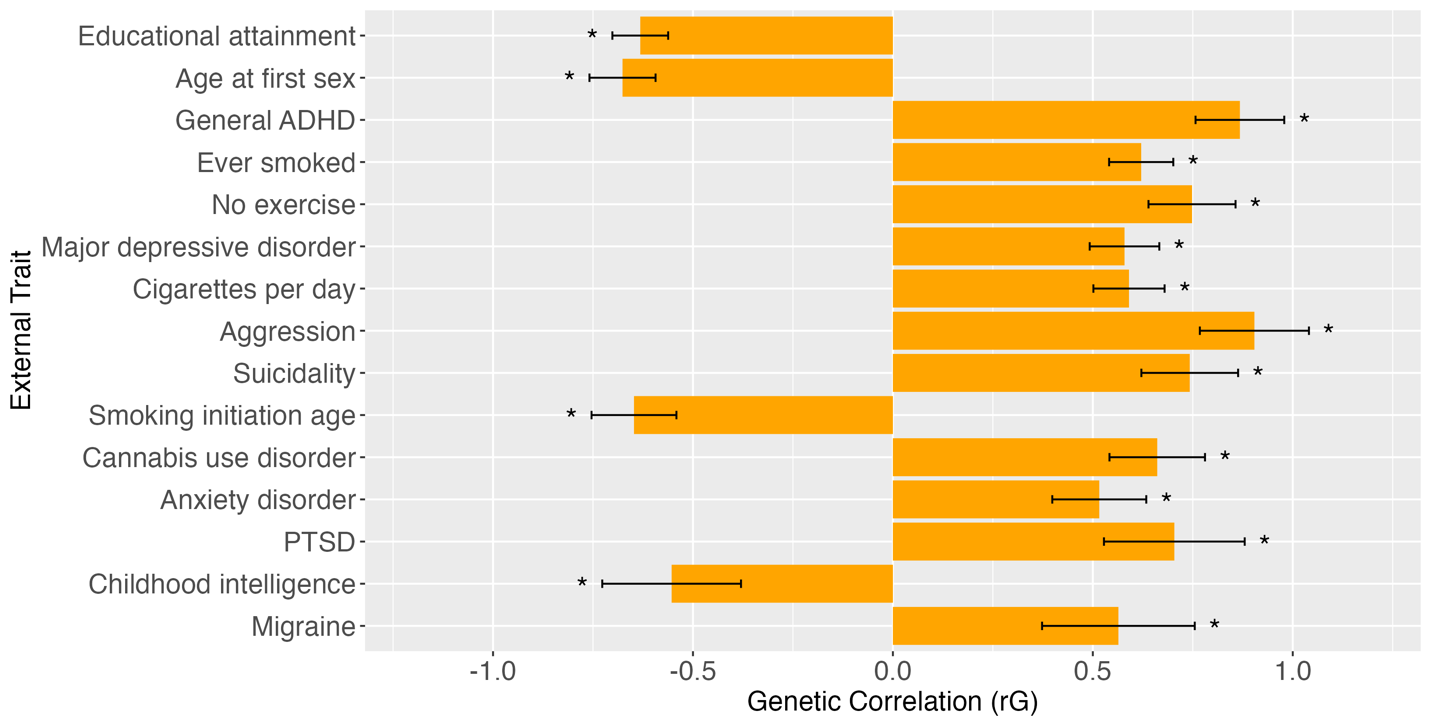

**Supplementary Figure 7. 15 most significant genetic correlations with adulthood diagnosed ADHD.** Sorted by smallest to largest p-value from top to bottom. Error bars display 1.96*Standard Error.

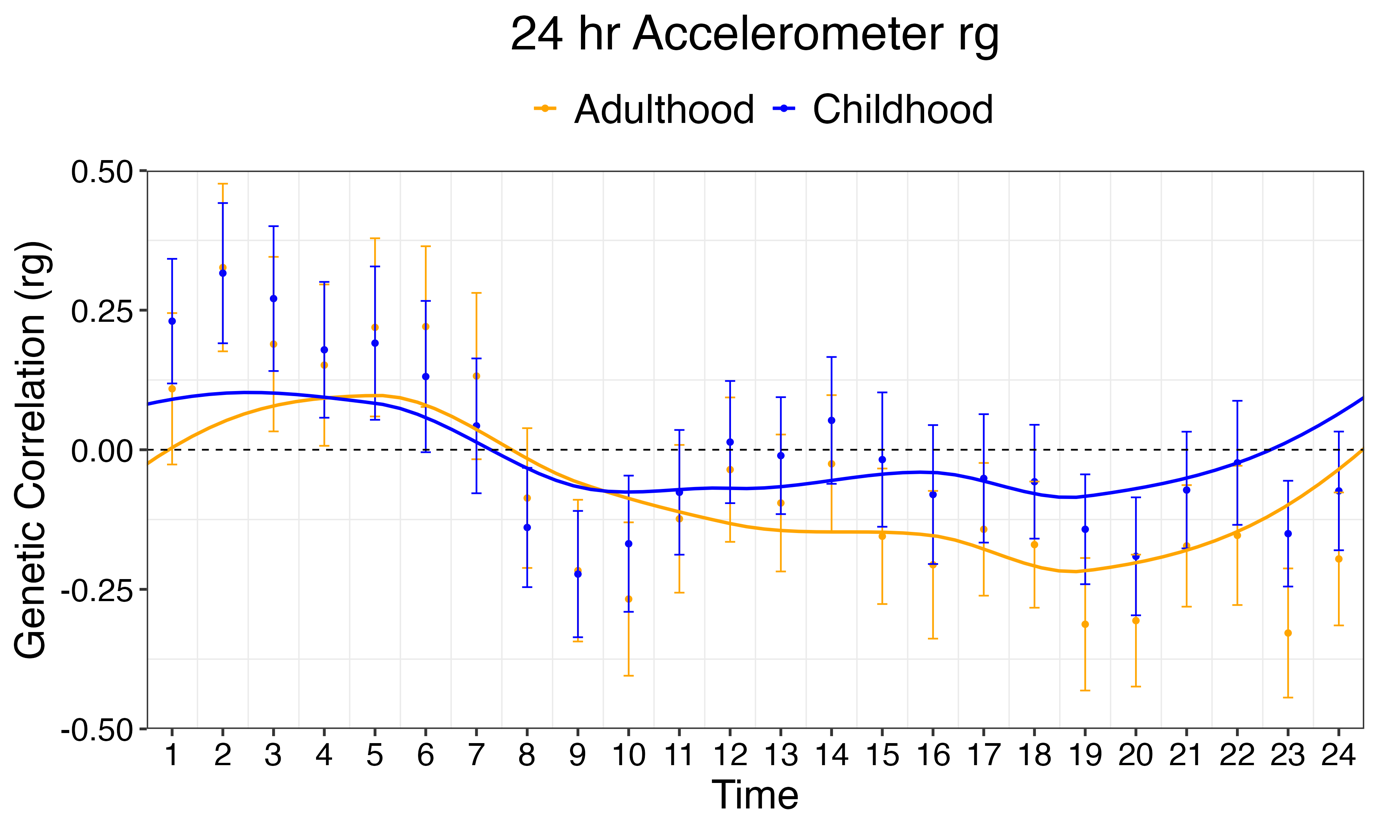

**Supplementary Figure 8. Genetic correlations between accelerometer based average total movement.** Accelerometer based movement within the 24h day beginning at midnight and childhood and adulthood diagnose ADHD. The error bars display 1.96*Standard Error for each time point.

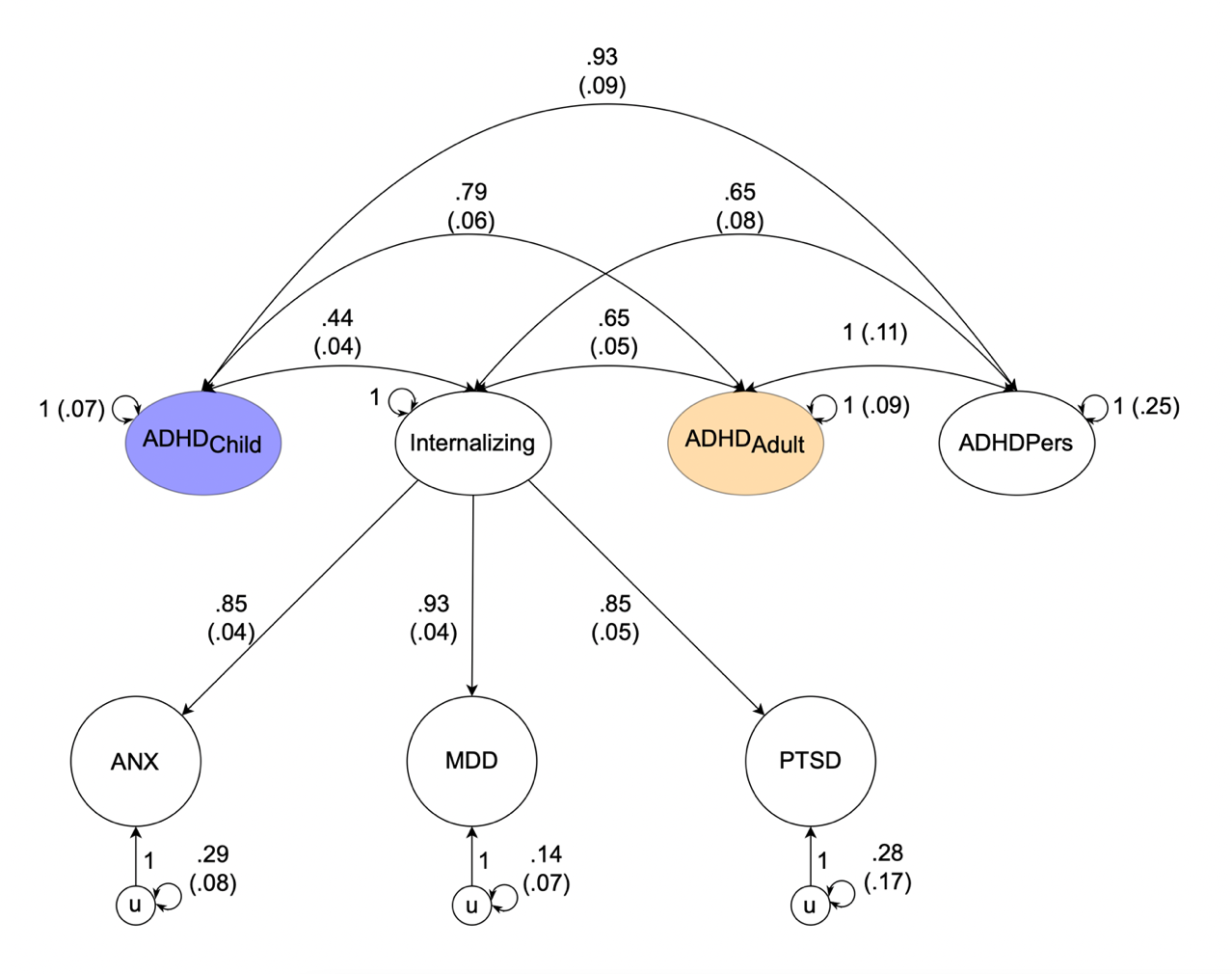

**Supplementary Figure 9. Path diagram of genetic correlations of adulthood, childhood, and persistent diagnosed ADHD with the internalizing factor.** Path diagram of the model used in Genomic SEM to test whether the genetic correlation between childhood or adulthood diagnosed ADHD with internalizing disorders could be constrained to be equal to that observed between persistent ADHD and internalizing. In this model internalizing is a common genetic factor of the genetic components of ANX, MDD, and PTSD and *u* is the residual genetic variance in these phenotypes, not explained by the internalizing factor. Latent variables are represented as circles. The genetic component of each phenotype is represented with a circle as the genetic component is a latent variable that is not directly measured but is inferred using LDSC. Single-headed arrows are regression relations, double-headed arrows connecting back to the same origin are variances, and double-headed arrows connecting two variables are correlations. Paths labeled 1 are fixed to 1. The variances of adulthood, childhood diagnosed, and persistent ADHD were constrained to be below 1 as these were otherwise estimated to be above 1. Additionally, we constrained the genetic correlation between persistent ADHD and adulthood diagnosed ADHD to be below 1, since it is otherwise estimated to be above 1. The low power of the persistent ADHD phenotype, and corresponding instability in model estimates, is one of the reasons we did not include the persistent ADHD phenotype in the main analyses. Results from this model should consequently be treated as preliminary and interpreted with caution.

### **Supplemental References**

1.	Bulik-Sullivan B, Finucane HK, Anttila V, et al. An atlas of genetic correlations across human diseases and traits. *Nat Genet*. 2015;47(11):1236-1241. doi:10.1038/ng.3406

2.	de la Fuente J, Davies G, Grotzinger AD, Tucker-Drob EM, Deary IJ. A general dimension of genetic sharing across diverse cognitive traits inferred from molecular data. *Nat Hum Behav*. 2021;5(1):49-58. doi:10.1038/s41562-020-00936-2

3.	Grotzinger AD, Fuente J de la, Privé F, Nivard MG, Tucker-Drob EM. Pervasive Downward Bias in Estimates of Liability-Scale Heritability in Genome-wide Association Study Meta-analysis: A Simple Solution. *Biological Psychiatry*. 2023;93(1):29-36. doi:10.1016/j.biopsych.2022.05.029

4.	Peyrot WJ, Boomsma DI, Penninx BWJH, Wray NR. Disease and Polygenic Architecture: Avoid Trio Design and Appropriately Account for Unscreened Control Subjects for Common Disease. *The American Journal of Human Genetics*. 2016;98(2):382-391. doi:10.1016/j.ajhg.2015.12.017
